# A Sex-Specific Genome-Wide Association Study of Depression Phenotypes in UK Biobank

**DOI:** 10.1101/2022.03.30.22273201

**Authors:** Patrícia Pelufo Silveira, Irina Pokhvisneva, David M Howard, Michael J. Meaney

## Abstract

**Background:** There are marked sex differences in the prevalence, phenotypic presentation and treatment response for major depression. While genome-wide association studies (GWAS) adjust for sex differences, to date no studies seek to identify sex-specific markers and pathways. In this study we performed a sex-stratified genome-wide association analysis for broad depression.

**Methods:** A genome-wide association study for broad depression was performed in the UK Biobank total participants (N=274,141), including only non-related participants, as well as separately in males (N=127,867) and females (N=146,274). Bioinformatics analyses were performed to characterize common and sex-specific markers and associated processes/pathways.

**Results:** We identified 11 loci passing genome level significance (P < 5* 10^−8^) in females and one in males. In both males and females, genetic correlations were significant between the broad depression GWA and other psychopathologies, however, correlations with educational attainment and metabolic features including body fat, waist circumference, waist-to-hip ratio and triglycerides were significant only in females. Gene-based analysis showed 147 genes significantly associated with broad depression in the total sample, 64 in the females and 53 in males. Gene-based analysis revealed “Regulation of Gene Expression” as a common biological process, but suggested sex-specific molecular mechanisms. Finally, sex-specific PRSs for broad depression outperformed total and the opposite sex PRSs in the prediction of broad MDD.

**Conclusions:** These findings provide evidence for sex-dependent genetic pathways for clinical depression as well as for health conditions comorbid with depression.

## Introduction

Major depressive disorder (MDD) is the leading contributor to the global burden of disease and disability worldwide (1–4). Depression shows marked sex differences with women significantly more affected in terms of prevalence, repeated occurrence, symptomatology and patterns of co-morbidity (5–12).

The neural correlates of MDD differ in males and females at the level of brain structure (13,14) and potentially cell-type composition (15). There are strikingly sex-specific transcriptional signatures of depression in corticolimbic brain regions associated with mood disorders (15–18).

These functional disparities are likely reflected in differences in the response to antidepressant treatment (19–25), which makes the understanding of sex-specific risk factors and vulnerabilities a pressing concern. An understanding of sex-specific pathways to psychopathology is also a fruitful approach to identifying novel mechanisms of pathophysiology (26).

Twin studies reveal evidence for moderate overall heritability for depression (27–29) with some evidence for greater heritability in women and for sex-specific genetic pathways to MDD (e.g. (28,30). Despite the evidence for sex-specific molecular mechanisms for depression, genome-wide association studies (GWASs) for MDD are still performed aggregating males and females into a single sample using sex as a covariate. The justification is based on the assumption that large sample sizes are essential and that the analyses of GWA datasets are adjusted by sex. There are two concerns with this approach: 1) pronounced sex differences in transcription suggest that aggregating males and females masks signals that are apparent only when considering datasets from males and females independently, and the noise of combining data regardless of sex might offset any advantage of a larger sample size; 2) collapsing male and female MDD data only permits the identification of molecular signals shared across males and females and thus to an incomplete understanding of the genetic risk factors linked to depression. The risk is that of ignoring critical, sex-specific molecular pathways and targets for the development of new therapies. The existing science suggests sex-specific approaches to treatment and is consistent with the broader objectives of precision medicine. However, to date this approach advances in the absence of an understanding of sex-specific genetic pathways.

Here we directly explored sex-dependency in the genetic architecture of MDD, by performing a follow-up analysis of a published GWAS for ‘broad depression’ (31) after stratifying the analysis by sex. Our findings are consistent with previous twin studies revealing sex-specific genetic architecture to clinical depression, and implicate sex-specific molecular pathways, aligned with genome-wide transcriptomic analyses (32).

## Methods and Materials

### Study population

The UK Biobank cohort is a population-based cohort consisting of 502,543 individuals aged 37-73 with genotyping data from 487,409 subjects. After retaining only Caucasians and variants with minor allele frequency <0.01 there were 276,511 individuals and 7,351,435 variants in the data set. The broad depression phenotype was defined according to Howard et al (31), resulting in 274,141 unrelated subjects with both the broad depression phenotype and genodata (see **Table S1-A** for case/control demographics). From the 132,066 subsample of related subjects, we selected one participant resulting in a test sample with 65,285 subjects (for details see **Supplemental Methods**). This research was conducted using the UK Biobank Resource under Application Number 41975. Approval for the UK Biobank was obtained by the North West Multicentre Research 580 Ethics Committee (REC reference 11/NW/0382), the National Information Governance Board for Health and Social Care and the Community Health Index Advisory Group. The raw data were used under license from UK Biobank (http://www.ukbiobank.ac.uk/).

### Association analysis

Linear regression analysis using BGENIE v1.132 was used to explore the effect of each SNP on the broad depression phenotype. Genetic correlations were calculated between the MDD phenotype for each sex and 235 other behavioral and disease related traits using LD Hub (33) (for details see **Supplemental Methods**).

### Gene-based analysis

Gene- and region-based analyses of the significant genes (P<2.6*10^−6^) were conducted using MAGMA (Multi-marker Analysis of GenoMic Annotation) available on FUMA_GWAS (Functional Mapping and Annotation of Genome-Wide Association Studies) (34). We compared male and female enrichment analyses using MetaCore™ (Clarivate Analytics, version 21.4). GTEx portal was used to identify brain expression quantitative loci (eQTL). Drug Targetor (35) was utilized to establish the potential mechanisms by which antidepressants act in male vs. female specific MDD. For details, see **Supplemental Methods**.

### Validation of the sex specific GWAS through polygenic risk scores

Aligned sample size GWAS were generated and used to calculate polygenic risk score (PRSs) at different p-value thresholds using PRSice software (36,37) for each subject of the test sample. Akaike information criterion (AIC) was utilized to identify the model that explains the greatest amount of variation using the fewest possible independent variables. Lower AIC scores associate with better-fitting models (for details see **Supplemental Methods**).

## Results

### Broad depression MDD GWAS in the total sample

A total of 18 independent loci showed genome-wide significance associated with broad depression (P<5*10^−8^) (**Data S1** and **Figure 1A**). The correlation of the beta coefficients between the Howard et al. (31) broad depression GWAS and our broad depression GWAS was r=0.91, and the correlation between the p values was r=0.72. There were 2,819 variants with P<10^−6^ for an association with broad depression for the total sample (**Figure 1A** and **Figure S1**). The phenotype examined did not show evidence of inflation of the test statistics due to population stratification, with any inflation due to polygenic signal (see **Table S2**). Genetic correlations from our total broad depression GWAS also replicated the findings from Howard et al. (31) **(Table 1** and **Data S2**). There were 24 significant correlations for broad depression with other traits (PFDR<0.05; **Data S2**). Correlations previously described in Howard et al. (31) between UK Biobank depression-related phenotypes and clinically defined MDD as well as schizophrenia (38) (rg=0.30) and bipolar disorder (39) (rg=0.34).

**Table 1.** Genetic correlations comparison between original Howard et al., 2018 Broad MDD GWAS, current study total sample GWAS, female specific and male specific Broad MDD GWAS.

|  | Broad MDD GWAS - Howard et al., 2018 |  |  |  | Broad MDD GWAS - total sample |  |  |  | Broad MDD GWAS - Females |  |  |  | Broad MDD GWAS - Males |  |  |  |
| --- | --- | --- | --- | --- | --- | --- | --- | --- | --- | --- | --- | --- | --- | --- | --- | --- |
| Trait | r<sub>g</sub> | SEM | P-value | FDR P-value | r<sub>g</sub> | SEM | P-value | FDR P-value | r<sub>g</sub> | SEM | P-value | FDR P-value | r<sub>g</sub> | SEM | P-value | FDR P-value |
| Age of first birth | -0.2981 | 0.0319 | 9.07E-21 | 2.67E-19 | -0.2616 | 0.0339 | 1.23E-14 | 2.63E-13 | -0.2751 | 0.0379 | 4.11E-13 | 1.07E-11 | -0.2238 | 0.0448 | 5.73E-07 | 1.22E-05 |
| Bipolar disorder | 0.3288 | 0.0366 | 2.53E-19 | 6.61E-18 | 0.3364 | 0.0381 | 1.16E-18 | 3.41E-17 | 0.2671 | 0.0466 | 9.62E-09 | 1.61E-07 | 0.3995 | 0.0559 | 8.84E-13 | 2.97E-11 |
| Body fat | 0.1782 | 0.0402 | 9.19E-06 | 1.20E-04 | 0.1719 |  | 1.96E-05 | 2.42E-04 | 0.2175 | 0.0485 | 7.32E-06 | 8.60E-05 | 0.0926 | 0.0469 | 4.84E-02 | 3.11E-01 |
| College completion | -0.2069 | 0.0384 | 6.83E-08 | 1.07E-06 | -0.2013 | 0.0405 | 6.72E-07 | 1.05E-05 | -0.2556 | 0.0413 | 6.11E-10 | 1.20E-08 | -0.1175 | 0.0613 | 5.54E-02 | 3.34E-01 |
| Depressive symptoms | 0.8268 | 0.0313 | 1.11E-153 | 2.61E-151 | 0.8415 | 0.0339 | 9.31E-136 | 1.09E-133 | 0.8197 | 0.0452 | 1.56E-73 | 1.83E-71 | 0.8424 | 0.0542 | 2.09E-54 | 2.46E-52 |
| Ever vs never smoked | 0.3161 | 0.046 | 6.56E-12 | 1.19E-10 | 0.2945 | 0.0481 | 8.82E-10 | 1.59E-08 | 0.3286 | 0.0566 | 6.39E-09 | 1.16E-07 | 0.2395 | 0.0576 | 3.16E-05 | 6.19E-04 |
| Excessive daytime sleepiness | 0.2408 | 0.043 | 2.10E-08 | 3.53E-07 | 0.2366 | 0.0458 | 2.34E-07 | 3.93E-06 | 0.2566 | 0.0556 | 3.88E-06 | 5.07E-05 | 0.2098 | 0.0597 | 4.00E-04 | 6.71E-03 |
| Insomnia | 0.4013 | 0.0462 | 3.82E-18 | 8.98E-17 | 0.4303 | 0.043 | 1.43E-23 | 4.80E-22 | 0.4658 | 0.0486 | 9.23E-22 | 5.42E-20 | 0.3764 | 0.0593 | 2.17E-10 | 5.67E-09 |
| Insomnia | -0.0876 | 0.0318 | 5.90E-03 | 3.85E-02 | 0.4085 | 0.0471 | 4.42E-18 | 1.15E-16 | 0.4463 | 0.0551 | 5.45E-16 | 1.83E-14 | 0.3507 | 0.0646 | 5.63E-08 | 1.32E-06 |
| Lung cancer | 0.2173 | 0.0544 | 6.37E-05 | 6.81E-04 | 0.214 | 0.0555 | 1.00E-04 | 9.79E-04 | 0.2684 | 0.066 | 4.80E-05 | 5.37E-04 | 0.1261 | 0.0689 | 6.73E-02 | 3.59E-01 |
| Major depressive disorder | 0.7856 | 0.0734 | 9.67E-27 | 3.79E-25 | 0.8441 | 0.0788 | 8.34E-27 | 3.92E-25 | 0.8077 | 0.086 | 6.20E-21 | 2.91E-19 | 0.8497 | 0.0984 | 5.69E-18 | 2.23E-16 |
| Neuroticism | 0.7266 | 0.0279 | 2.25E-149 | 2.64E-147 | 0.7414 | 0.0292 | 1.30E-142 | 3.06E-140 | 0.749 | 0.035 | 1.66E-101 | 3.90E-99 | 0.7181 | 0.0419 | 5.55E-66 | 1.30E-63 |
| Neuroticism | 0.8204 | 0.0965 | 1.92E-17 | 4.09E-16 | 0.855 | 0.0997 | 9.78E-18 | 2.30E-16 | 0.8968 | 0.1209 | 1.19E-13 | 3.50E-12 | 0.776 | 0.1089 | 1.04E-12 | 3.06E-11 |
| Number of children ever born | 0.1795 | 0.0409 | 1.16E-05 | 1.44E-04 | 0.1734 | 0.0438 | 7.50E-05 | 8.01E-04 | 0.1974 | 0.0494 | 6.41E-05 | 6.85E-04 | 0.1347 | 0.056 | 1.61E-02 | 1.58E-01 |
| PGC cross-disorder analysis | 0.4712 | 0.0372 | 1.04E-36 | 6.14E-35 | 0.4839 | 0.0404 | 4.13E-33 | 2.43E-31 | 0.4125 | 0.0476 | 4.20E-18 | 1.64E-16 | 0.5514 | 0.0587 | 5.47E-21 | 2.57E-19 |
| Schizophrenia | 0.3008 | 0.0285 | 5.28E-26 | 1.77E-24 | 0.3026 | 0.0302 | 1.12E-23 | 4.39E-22 | 0.2299 | 0.0331 | 3.53E-12 | 8.30E-11 | 0.3806 | 0.0401 | 2.48E-21 | 1.46E-19 |
| Subjective well being | -0.6069 | 0.0406 | 1.33E-50 | 1.04E-48 | -0.6228 | 0.0415 | 6.49E-51 | 5.08E-49 | -0.5937 | 0.0492 | 1.77E-33 | 1.39E-31 | -0.6338 | 0.0583 | 1.54E-27 | 1.21E-25 |
| Triglycerides | 0.1183 | 0.0303 | 9.17E-05 | 9.37E-04 | 0.1318 | 0.0307 | 1.73E-05 | 2.26E-04 | 0.1529 | 0.039 | 8.81E-05 | 9.00E-04 | 0.0979 | 0.0397 | 1.37E-02 | 1.46E-01 |
| Waist-to-hip ratio | 0.1211 | 0.0282 | 1.77E-05 | 1.98E-04 | 0.1183 | 0.0284 | 3.12E-05 | 3.67E-04 | 0.1531 | 0.0337 | 5.46E-06 | 6.75E-05 | 0.0603 | 0.0373 | 1.06E-01 | 4.46E-01 |
| Years of schooling (proxy cognitive performance) | -0.1836 | 0.037 | 7.15E-07 | 1.05E-05 | -0.1776 | 0.0384 | 3.72E-06 | 5.14E-05 | -0.2081 | 0.0411 | 4.17E-07 | 6.53E-06 | -0.1227 | 0.0544 | 2.41E-02 | 2.18E-01 |
| Years of schooling 2013 | -0.1768 | 0.0373 | 2.16E-06 | 2.99E-05 | -0.1762 | 0.0379 | 3.36E-06 | 4.94E-05 | -0.2067 | 0.0414 | 5.81E-07 | 8.53E-06 | -0.1192 | 0.0539 | 2.71E-02 | 2.20E-01 |
| Years of schooling 2016 | -0.1703 | 0.0223 | 2.08E-14 | 4.08E-13 | -0.1613 | 0.0229 | 1.95E-12 | 3.82E-11 | -0.1901 | 0.0284 | 2.09E-11 | 4.47E-10 | -0.1139 | 0.0305 | 2.00E-04 | 3.62E-03 |
| Coronary artery disease | 0.1236 | 0.0288 | 1.71E-05 | 1.98E-04 | 0.122 | 0.0304 | 5.90E-05 | 6.60E-04 | 0.1356 | 0.0375 | 3.00E-04 | 2.71E-03 | 0.1004 | 0.0407 | 1.36E-02 | 1.46E-01 |
| Inflammatory Bowel Disease (Euro) | 0.4379 | 0.0393 | 7.63E-29 | 3.59E-27 | 0.132 | 0.0345 | 1.00E-04 | 9.79E-04 | 0.1275 | 0.0406 | 1.70E-03 | 1.43E-02 | 0.142 | 0.0433 | 1.00E-03 | 1.57E-02 |
| Lung cancer (all) | 0.2122 | 0.0605 | 5.00E-04 | 4.35E-03 | 0.2121 | 0.0617 | 6.00E-04 | 5.42E-03 | 0.2826 | 0.0724 | 9.54E-05 | 9.34E-04 | 0.1 | 0.0779 | 1.99E-01 | 5.30E-01 |
| Squamous cell lung cancer | 0.3099 | 0.0907 | 6.00E-04 | 5.04E-03 | 0.3001 | 0.0911 | 1.00E-03 | 8.39E-03 | 0.3799 | 0.1018 | 2.00E-04 | 1.88E-03 | 0.1613 | 0.1163 | 1.65E-01 | 5.04E-01 |
| Waist circumference | 0.1019 | 0.0277 | 2.00E-04 | 1.88E-03 | 0.1025 | 0.0297 | 6.00E-04 | 5.42E-03 | 0.1627 | 0.0346 | 2.62E-06 | 3.62E-05 | 0.0064 | 0.0385 | 8.69E-01 | 9.26E-01 |

**Figure 1.**
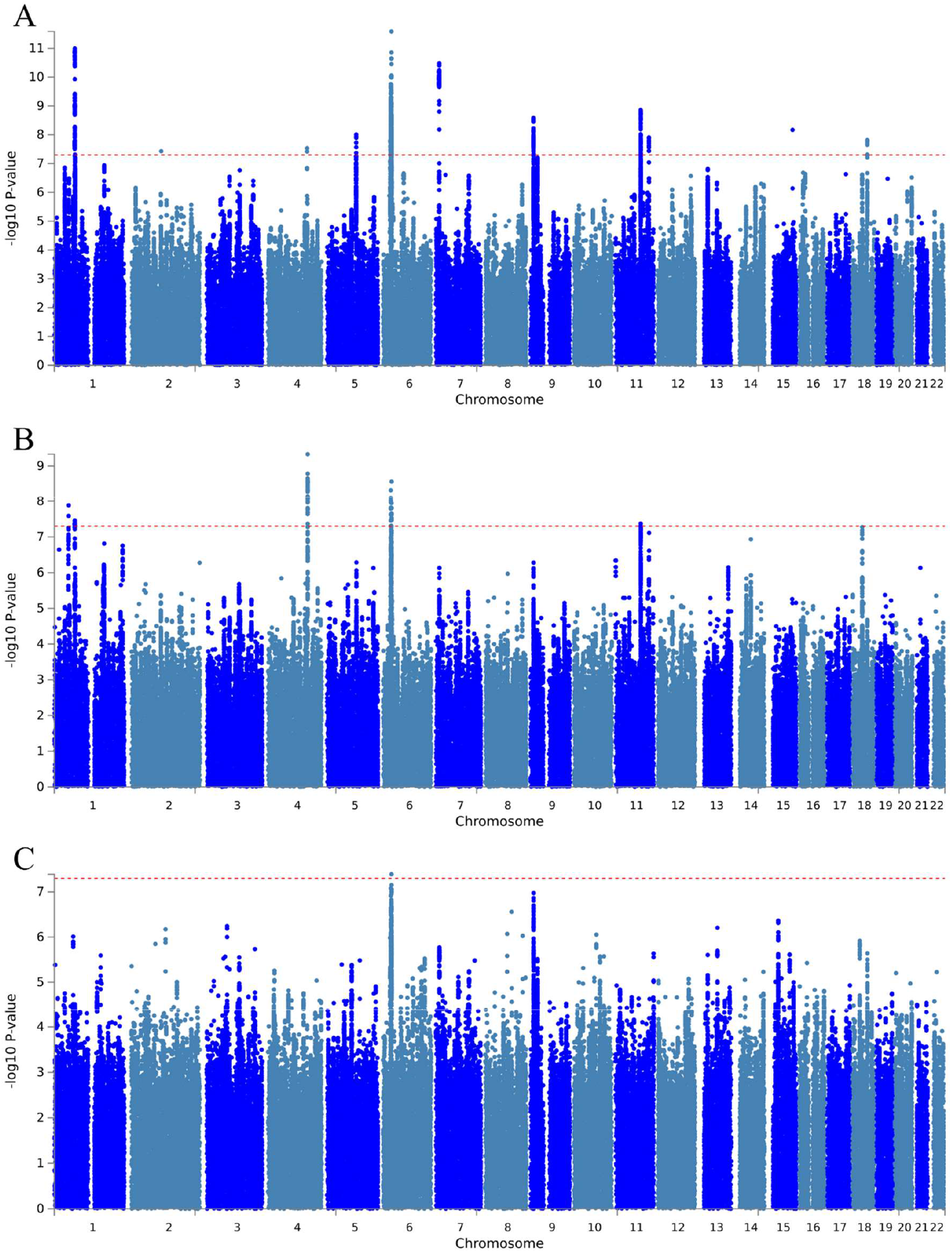
Manhattan plot of all the variants analyzed in UK Biobank for broad depression in the **(A)** total sample (N=274,141), **(B)** females (N=146,274), and **(C)** males (N=127,867).

### Sex-specific broad depression MDD GWAS

We then stratified our total sample by sex to perform sex-specific GWASs separately for 127,867 non-related male and 146,274 non-related female participants (see **Table S1-B and C** for case/control demographics). Results for the associated variants with broad depression phenotype in males and females are provided in **Data S1, Figures 1B** and **S2** for females and **1C** and **S3** for males. Broad depression in females and males did not show evidence of inflation of the test statistics due to population stratification, with any inflation due to polygenic signal (**Table S2**). Eleven loci passed genome level significance (P<5*10^−8^) in females and one in males. Interestingly, among these loci was a SNP (rs10501696) that mapped to *GRM5* reported in Howard et al. (31), significant only in females.

### Sex-dependent genetic correlations

Depression is comorbid with a range of other diseases including conditions considered to be not primarily of brain origin, especially cardio-metabolic conditions. Our findings (**Table 1**) demonstrated 25 and 13 significant (PFDR<0.05) correlations with other traits in females and males, respectively. Genetic correlations between the UK Biobank depression-related phenotypes and clinically defined MDD, schizophrenia (38), bipolar disorder (39), neuroticism (40), subjective well-being (40), PGC cross-disorder (41), insomnia (42,43) and smoking (44) were significant for both females and males.

Two phenotypic categories showed striking, sex-dependent correlations, each with greater evidence for associations amongst females (**Table 1**). While analyses with both male and female participants showed associations with measures of academic achievement, the evidence for significant genetic correlations was stronger and more pervasive in females. Genetic significant (PFDR<0.05) correlations only in females included multiple analyses of years of schooling and college completion (rg=-0.21 and -0.26, respectively) (45). The most striking sex difference were those between broad MDD and GWASs for metabolic features, including body fat (46), waist circumference (47), waist-to-hip ratio (47) and triglycerides (48), significant at PFDR<0.05 only in females.

### Sex-specific broad MDD GWAS: Gene-based enrichment analysis

SNPs with P<10^−5^ were selected to compare female and male GWAs. 147 genes were significantly associated with broad depression in the total sample, 64 in females and 53 in males (**Figures S4, S5 and S6, Data S3A-C**). In addition to *GRM5, ELAVL4*, implicated in depression and in epigenome-wide association studies of suicide (49), was significant in females, but not males. In contrast, 29 genes were significant only in males. Notably, *TCF4, NKAPL, ZKSCAN3, ZSCAN16, ZSCAN31* have been associated with depression in previous GWASs (50–54). Twenty-four common genes were found when comparing the list of total, males and females broad MDD GWAS, which were enriched for biological processes related to sensory perception and G protein-coupled receptor signaling pathways.

There were several Pathway Maps in common between genes derived from the male and female broad MDD GWASs, the most significant of which is glutathione metabolism (PFDR<0.025) (**Data S4A**). Glutathione is involved in antioxidant defense and regulation of gene expression, cell proliferation and apoptosis, signal transduction, and immune response (55). Several common GO processes were found between males and females, the most significant of which were negative regulation of viral life cycle, regulation of viral release from host cell, regulation of transcription DNA-templated, regulation of nucleic acid-templated transcription, regulation of RNA biosynthetic process (all PFDR<0.002) (**Data S4B**). Common genes between males and females were enriched for diseases related to neurocognitive and neurodegenerative conditions (**Data S4C**).

Enrichment analysis for the genes uniquely identified in male- or female-specific broad MDD GWAS’s is provided in **Data S5A-E**. Male-specific broad MDD GWAS genes were enriched for several Pathway Maps, GO processes and process networks (**Data S5A, B, D**) with epigenetic regulation of gene expression as the recurrently enriched pathway. Female-specific broad MDD GWAS genes did not show significant enrichment for Pathway Maps but, as in males, were enriched for regulation of gene expression (PFDR<0.01, **Data S5C**).

It is noteworthy that “regulation of gene expression” was the most significant common GO process associated with genes from both male- and female-specific broad MDD GWAS, but this finding was due to sex-specific gene networks (**Figure 2**). In males, “regulation of gene expression” was mapped to genes including *TCF4* as well as an impressive number of genes coding for histone protein variants. *TCF4* is a known regulator of epigenetic states including DNA methylation (56). In females, “regulation of gene expression” was associated with a number of neurexin-related genes, *DRD2* and *GRM5* genes.

**Figure 2.**
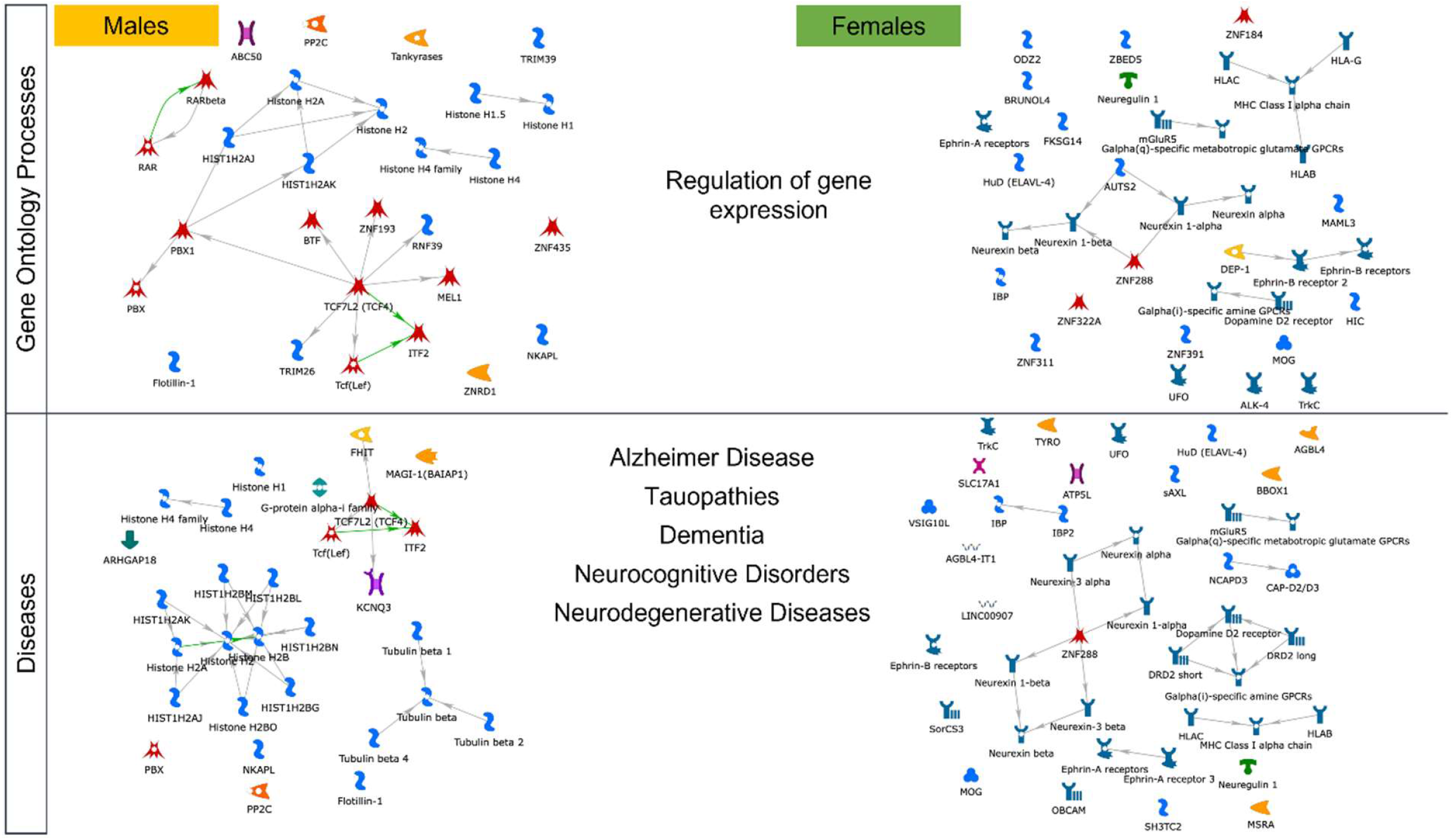
Enrichment analysis of genes associated with male-specific or female specific broad MDD GWAS. Processes and diseases are commonly enriched in both sexes, but due to unique, sex-specific mechanisms.

Several psychopathology and neurodegenerative disease terms were associated with genes from both male- and female-specific broad MDD GWAS (**Figure 2**), but this finding was due to sex-specific gene networks. In males, brain pathology was associated with genes involved in epigenetic processes and regulation of neurotransmitter release. In females, brain pathology was related to *DRD2* signaling, and an important number of genes related to adaptive immunity. Taken together, the findings depicted in **Figure 2** suggest that MDD-related alterations in gene expression regulation and brain pathology in males and females occur via unique, sex-specific mechanisms.

### Sex-specific broad MDD GWAS: eQTL’s, transcription factors and drug targets

We sought eQTLs among the SNPs identified in male- and female-specific broad MDD GWAS (top 125 significant SNPs, P <10^−7^ in females and P <9.16*10^−7^ in males). There was a striking sex difference with almost twice the percentage of SNP/eQTLs in males [18% compared with females 10% (χ2 = 945.9, p<0.00001)] considering all brain regions together.

The cerebellum showed a high number of eQTLs in both male- and female-specific GWASs (**Figure S7**). There was a highly sex-specific distribution of identified eQTLs across brain regions. eQTLs from the female-specific GWAs in the caudate-basal ganglia (χ2 = 38.8; p<.00001), putamen-basal ganglia (χ2 = 31.6; p<.00001), and hippocampus (χ2 = 19.7; p<.00001) significantly exceeded that derived from the male-specific GWAs. The most notable finding was that of increased eQTLs in the basal ganglia of females, including the ventral striatum/nucleus accumbens, which are prominent dopamine target regions.

A conspicuous pattern apparent in both males and females was the enrichment for transcription factors linked to immune responses and NFK-ß signaling, including FOXP3, RBPJ kappa, RUNX1 and TAL1. BMAL1, a critical regulator of circadian rhythms (57), was highly enriched amongst the genes from both the male- and female-specific GWASs (**Data S6A-B**). This finding is interesting considering the highly significant genetic correlation between both the male- and female-specific GWASs for broad MDD and that for insomnia (see **Table 1**).

Transcription factors uniquely associated with genes identified in the female broad MDD GWAS were linked to oxidative stress, apoptosis and type II diabetes (e.g. HNF3-beta, MafA, p63, RelA, VDR), which aligns with the genetic correlation of the female-specific GWAS with cardio-metabolic conditions (see **Table 1**). Transcription factors linked specifically with genes identified in the male-specific broad MDD GWAS were related to tissue and neuron differentiation as well as epigenetic processes (e.g. E2F1, Esrrb, NRSF, ZFX, ZNF423), which is consistent with the results of the gene enrichment analyses (see **Figure 2**) that underscore the potential role for chromatin remodeling factors in males.

The central biological processes associated with broad MDD and targeted by antidepressants were strongly associated with: (1) epigenetic processes such as chromatin assembly, cell cycle regulation as well as inflammation (mainly through IL-7), in males; (2) neuronal migration, regulation of neurotrophic factors and synaptic plasticity, and dopamine neurotransmission, in females (**Data S7A-B**). The implication of the dopamine neurotransmission is consistent with the pathways identified in females in the gene enrichment analyses (**Figure 2**), likewise, the finding of chromatin assembly in males, a process intimately linked to histone proteins. These findings suggest that the biological processes targeted by antidepressants and involved in genetic architecture of MDD differ in males and females, implying sex-dependent therapeutic pathways.

### Validation of the sex-specific broad MDD GWAS using polygenic risk scores

We calculated PRSs using our total, male- and female-specific GWASs in a UK Biobank test sample of 65,285 non-related individuals (**Table S3**). In males, the male-specific PRS showed better predictive value for the broad MDD outcome (**Figure 3B**) than did the total PRS derived from a similar sized GWAS (**Figure 3C**) or the female-specific PRS (**Figure 3A**) (AICm=33962.86, AICf=34001.21, AICt=33965.37). Similarly, the PRS derived from the female-specific GWAS showed better predictive value for the broad depression outcome among females (**Figure 3E**) than did the total PRS from a GWAS of comparable size (**Figure 3G**) and male-specific PRS (**Figure 3F**) (AICf=48987.21, AICm=49100.48, AICt=49016.82). The well-established effect of GWAS sample size to best predict the outcomes was observed, as the combined, mixed GWAS sample had a similar predictive capacity to the sex-specific male or female-specific GWAS (**Figure 3D and 3H**). Although a larger GWAS does indeed have a similar predictive capacity than the sex-specific GWAS, it does not have the ability to disentangle sex-specific mechanisms and drug targets.

**Figure 3.**
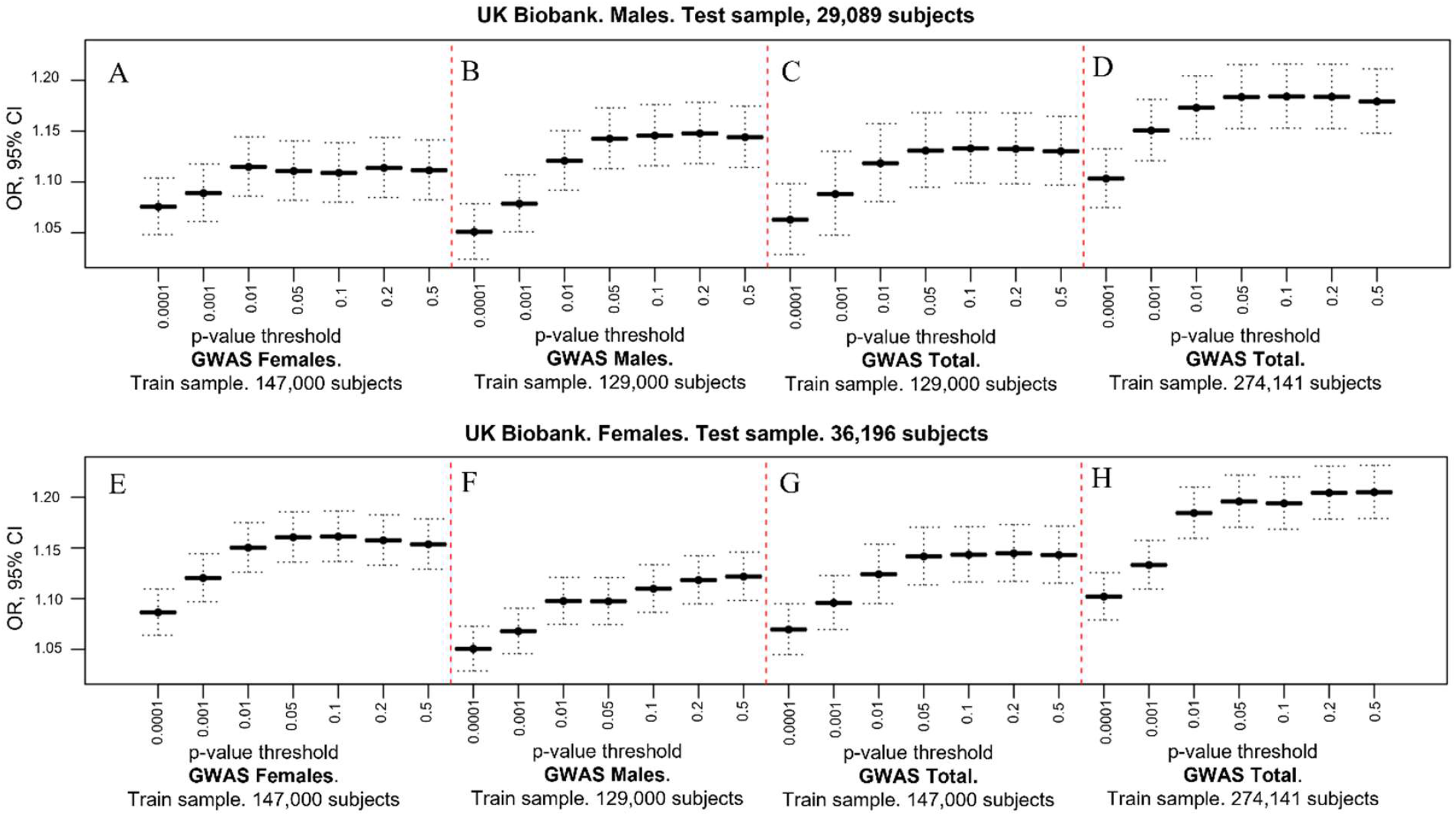
Comparison between odds ratios (ORs) of associations between the polygenic risk scores (PRS) for Broad MDD in a test sample of males (A-D) and females (E-H) using the male-specific broad MDD GWAS (M129), female-specific broad MDD GWAS (F147), or a mixed sample MDD GWAS of similar size (T129 or T147, respectively). The ORs of associations using the total mixed sample MDD GWAS (T276) is shown in panels D for males participants and panel H for female participants.

## Discussion

Sex differences in the prevalence and severity of depression are well established. Nevertheless, research to elucidate the genetic architecture of depression has not been designed to examine sex-specific pathways (58). The current study addresses this gap. To our knowledge our results are the first extensive molecular analysis of sex-specific genetic pathways for MDD. Our findings reveal evidence for sex-differences in the genetic pathways to MDD and are consistent with previous, genetically-informed epidemiological studies (27,28) suggesting that the influences of genetic variants on the risk for depression are sex-specific. We showed that a PRS derived from a female-specific, genome-wide analysis outperformed a version derived from a male-specific analysis in the prediction of broad MDD in females; the reverse was true for the prediction of broad MDD in males (**Figure 3**). While the gene enrichment analysis identified “regulation of gene expression” as a common biological process linking genetic variants to broad MDD, the underlying gene pathways were highly sex-dependent (**Figure 2**). Regulation of gene expression in males was associated with epigenetic mechanisms, notably variants in genes coding for the core histone proteins. In contrast, regulation of gene expression in females associated with neurexin as well as the DRD2 and mGluR5 receptors.

The implication of the DRD2 receptor in the “regulation of gene expression” pathway is consistent with previous reports. In their MDD GWAS Howard et al. (31) explored drug – gene interactions and identified the DRD2 receptor as the primary target. Levey et al. (59) mapped genes from their GWAS meta-analysis of depression to expression QTL data in GTEx revealing a transcriptome-wide association with a predicted decrease in nucleus accumbens DRD2 expression. Our findings suggest that these results might be largely driven by data from women. Note the inclusion of the DRD2 receptor in the female-specific “Regulation of Gene Expression Pathway” (**Figure 2**). Our gene-based eQTL analysis of the female-specific GWAS revealed an enrichment for eQTL’s in the caudate, which includes the dorsal and ventral striatum/nucleus accumbens, brain regions rich in DRD2 receptors.

Considerable evidence from both pre-clinical and clinical studies show the importance of dopaminergic projections from the ventral tegmental area (VTA) to nucleus accumbens for depression (60). Anhedonia is a core feature of depression and the mesolimbic dopamine pathway mediates activation of the reward system (61). Deep brain stimulation affecting the nucleus accumbens has sustained efficacy in treatment-resistant depression (62). The mesolimbic dopamine pathway is critical for chronic stress-induced depressive-like behaviors in rodents (63–65). Buproprion, a mixed norepinephrine/dopamine-reuptake inhibitor, is an accepted treatment for depression with efficacy comparable to that of SSRI’s (66,67). Pre-clinical studies reveal greater DRD2-mediated reward enhancing effects of buproprion in females than males (68). Rodent studies consistently show sex differences in dopaminergic systems in relation to reward processing and activation of behavioral responses to stress (69). Human PET imaging studies using [11C]raclopride, a DRD2/3-specific ligand, show in vivo evidence for stress-induced DRD2 signaling in the ventral striatum (70) and decreased ventral striatal D2 synaptic activity in depressed patients (71). Human neuroimaging studies reveal greater stress-induced activation of the ventral striatum in women compared to men (72,73).

Eleven loci passed genome level significance (P<5*10^−8^) in females including a SNP (rs10501696) mapped to *GRM5* also reported in Howard et al. (31). Wray et al. (50) reported on a meta-analysis of seven independent cohorts and identified a significant association of *GRM5* with MDD. *GRM5* gene, which encodes the mGluR5 metabotropic glutamate receptor, was also featured in the female-specific “Regulation of Gene Expression Pathway” (**Figure 2**). These sex-specific *GRM5* findings are consistent with Gray et al. (74) reporting extensive sex-dependent differences in glutamate receptor gene expression in post-mortem samples from MDD and control subjects, and higher expression levels of *GRIN1, GRIN2A-D, GRIA2-4, GRIK1-2, GRM1, GRM4, GRM5* and *GRM7* in female MDD patients; amongst males only *GRM5* expression differed and in the opposite direction from females.

The G-protein coupled mGluR5 receptor is located both pre- and post-synaptically, regulating synaptic plasticity (75). mGluR5-/- mice show increased stress-induced depressive-like behaviors (76). Reductions in mGluR5 protein levels are apparent in multiple rodent models of depression (77,78), in contrast to the increases in mGluR5 levels observed following anti-depressant treatments (79). *In vivo* PET imaging with [11C]ABP688 reveals lower mGluR5 receptor density in MDD in several regions (80), which seems to be associated with response to antidepressant treatment (81).

Our gene level analysis identified “regulation of gene expression” as the primary biological process for both males and females, revealing the involvement of neurexin signaling in females (**Figure 2**), which is critical for the formation of neural circuits and implicated in a range of neuropsychiatric disorders (82,83). To our knowledge previous GWASs have not provided evidence for an association between polymorphisms in neurexin genes and MDD. However, alternative gene sequencing platforms do suggest a potential role for neurexin in MDD. Rucker et al. (84) found significant enrichment of genomic and exonic deletion CNVs in cases of recurrent depression. The analysis showed overlap with CNVs previously associated with schizophrenia, including neurexin 1. This same exonic *NRXN1* deletion CNV was also associated with a poor treatment response to antidepressants (85). An intronic SNP in neurexin 3 was significantly associated with symptom improvement following citalopram/escitalopram treatment (86). Transcriptomic analyses with post-mortem human brain samples reveal significant sex differences in both the expression and splicing of neurexin genes (87,88). Studies with model systems suggest a prominent role for neurexin in establishing and maintaining sexually-dimorphic neuronal circuits (89).

The “Regulation of Gene Expression” process in males was associated with histone modifications. An analysis from the Psychiatric Genomics Consortium (90) examined common pathways across GWASs for schizophrenia, major depression and bipolar disorder, identifying histone methylation as the strongest emerging common process. Pre-clinical models of depression underscore the importance of histone methylation (91,92), with emerging evidence from human post-mortem analyses (93). Our analysis identified genes coding for actual histone protein variants rather than enzymes associated with methylation. Histone protein variants emerge from a histone gene cluster as key components of the transcriptional machinery (94). Histone variants affect nucleosome dynamics associated with activity-dependent transcription in brain (95,96) and differ in the capacity for post-translational modification. Hodes et al. (97) summarized the existing evidence for sex-dependent effects of epigenetic mechanisms in rodent models of depression. The extensive sex-dependent transcriptomic profiles of depression together with the sex differences in eQTLs described here points to an important area for future analyses.

While we emphasize the sex-dependent features of our dataset, there were important points of convergence between male and female analyses. Our transcription factor enrichment analysis revealed common transcriptional signals, most notably factors involved in NFK-ß-signaling such as FOXP3, TAL1 and RUNX1, consistent with the proposed association between inflammation and MDD (98). Another common factor was BMAL1, a regulator of circadian rhythms and sleep. Genetic correlation between the broad MDD GWAS and insomnia was significant in both sexes (**Table 1**). Notably absent in the transcription factor enrichment analysis were sex-steroid receptors, which serve as ligand-gated regulators of gene expression.

Genetic correlations for male- and female-specific GWASs revealed significant associations with previous GWASs for a range of psychiatric disorders (see **Table 1**). Nevertheless, a striking sex difference in the genetic correlations were those between broad MDD and GWASs for metabolic features, including body fat, waist circumference, waist-to-hip ratio and triglycerides, all of which associate with an increased risk for cardio-metabolic disease and were highly correlated in women, not men (**Table 1**). Co-morbidity between MDD and cardio-metabolic diseases is well established (99) being significantly more prevalent in women (100–103) and see (104) for a review]. Marcus et al. (105) reported that women with MDD were more likely to endorse changes in appetite and weight gain than were men in the STAR*D (www.star-d.org) trial. Our findings suggest that sex-specific genetic pathways may explain, in part, this increased co-morbidity in women. The sex-specific MDD pathway analyses highlighted dopamine signaling in females and analyses of the sex-specific GWAS’s showed greater eQTLs in the basal ganglia in women, including the caudate and nucleus accumbens. Dopamine signaling through DRD2 receptors in these regions regulates appetite and feeding behavior (106).

Our study was an exploratory investigation into potential sex-dependent genetic pathways to depression. Our genome-wide analyses were not sufficiently powered to provide definitive identification of sex-dependent loci associated with MDD, requiring extension with sex-based stratification of larger sample sizes and diverse populations. Nevertheless, we note that despite the comparatively smaller sample size, the female-specific GWAS did yield 11 loci that passed genome-wide statistical significance. A comparably powered analysis failed to yield significant loci in the male-specific GWAS. Sex-stratified analyses with larger sample sizes are required to examine whether this reflects a greater genetic contribution to MDD in women.

The reliance on a self-reported, “broad MDD” designation is a limitation of this exploratory study. However, Howard et al. (31) showed a highly significant genetic correlation (rG= 0.86) between broad depression and clinically diagnosed MDD [also see (107)]. The analyses are also limited by the focus on British Caucasians, with subjects generally exceeding the health and wealth of the overall British population.

In summary, our exploratory study suggests that the genetic background linked to human major depression includes sex specific variants. There are both common and unique biological mechanisms mapped from male-specific and female-specific broad MDD GWAS. Common processes like regulation of gene expression and diseases like brain pathology emerged from sex-specific gene networks. Our findings may contribute for the development of tailored therapeutic options. The consideration of sex-specific molecular alterations related to major depression during disease management can lead to more effective response to antidepressants. Significantly larger samples across more diverse populations will be required to meet this objective. Our results are intended to add to the rationale for studies of sex-specific mechanisms.

## Data Availability

All data produced in the present study are available upon reasonable request to the authors and with permission of UK Biobank.

## Acknowledgements

We are grateful to UK Biobank participants. This research has been conducted using the UK Biobank Resource under application number 41975. This research was funded by the Hope for Depression Research Foundation (MJM).

## Disclosures

The authors report no biomedical financial interests or potential conflicts of interest.

## Notes

### Competing Interest Statement

The authors have declared no competing interest.

### Funding Statement

This study was funded by the Hope for Depression Research Foundation.

### Author Declarations

Approval for the UK Biobank was obtained by the North West Multicentre Research 580 Ethics Committee (REC reference 11/NW/0382), the National Information Governance Board for Health and Social Care and the Community Health Index Advisory Group.

