## Supplemental Methods for "A Sex-Specific Genome-Wide Association Study of Depression Phenotypes in UK Biobank"

**Supplement - Methods and Materials**

**Study population**

The UK Biobank cohort is a population-based cohort consisting of 502,543 individuals aged 37-73 recruited at 23 centers across the United Kingdom between 2006 and 2010. Participants provided both phenodata and genodata. Genotyping data was available for 487,409 subjects. We excluded participants who withdrew their consent, with inconsistencies in genetic and reported sex, as well as outliers for heterozygosity. Also, we retained only those subjects who identified themselves as “Caucasians”. After applying the above-mentioned criteria, there were 408,577 subjects. Next we excluded 132,066 participants with shared relatedness of up to the third degree (kinship coefficients >0.044 calculated using the KING software). We also removed variants with minor allele frequency <0.01, an imputation accuracy Info score < 0.1 as well as duplicated and ambiguous SNPs. As a result, there were 276,511 individuals and 7,351,435 variants in the data set. The broad depression phenotype was defined according to Howard et al. (1), which resulted in 113,769 cases and 208,811 controls (total = 322,580, prevalence = 35.27%). There were 274,141 unrelated subjects with both the broad depression phenotype and genodata. From the subsample of related subjects (132,066 participants) we selected one participant per each group of related participants (genetic relatedness < 0.025) based on genomic relationship matrix (calculated using Genome-wide Complex Trait Analysis (GCTA 1.93.2), which resulted in 65,285 subjects with the broad depression phenotype (test sample). This research was conducted using the UK Biobank Resource under Application Number 41975. Approval for the UK Biobank was obtained by the North West Multicentre Research 580 Ethics Committee (REC reference 11/NW/0382; www.ukbiobank.ac.uk/ethics/), the National Information Governance Board for Health and Social Care and the Community Health Index Advisory Group.

**Data availability**

The raw genetic and phenotypic data that support the findings of this study are available from UK Biobank but restrictions apply to the availability of these data, which were used under license for the current study, and so are not publicly available. Data are, however, available from the authors upon reasonable request and with permission of UK Biobank (http://www.ukbiobank.ac.uk/).

**Association analysis**

We applied linear regression analysis using BGENIE v1.132 to explore the effect of each SNP on the broad depression phenotype. Before performing the regression analysis, we adjusted the outcome for sex, age, genotyping array and eight genetic principal components. Linkage Disequilibrium Score regression (LDSR) (2,3) was used to determine whether there was elevation of the polygenic signal due to population stratification, by examining the intercept for evidence of significant deviation (± 1.96 standard error) from 1. The genomic inflation factor (λGC) was also reported for each phenotype. Genetic correlations were calculated between the MDD phenotype for each sex and 237 other behavioural and disease related traits using LD Hub (2). P-values were false discovery rate (FDR) adjusted using the Benjamini and Hochberg approach.

**Gene-based analysis**

Gene- and region-based analyses of the significant genes (P < 2.6*10^-6^) were conducted using MAGMA (Multi-marker Analysis of GenoMic Annotation) available on FUMA_GWAS (Functional Mapping and Annotation of Genome-Wide Association Studies) (4). For gene-set pathway analysis we used the results obtained from the gene-based analysis considering SNPs at 10^-5^ as the threshold to conduct a further gene-set pathway analysis to test for gene enrichment using FUMA_GWAS (Functional Mapping and Annotation of Genome-Wide Association Studies), Gene2func, gene set analysis, GO molecular functions. We also compared male and female enrichment analyses using MetaCore™ (Clarivate Analytics, version 21.4) (https://portal.genego.com). We used “Particular set” sorting method in MetaCore and exported significant common elements (FDR < 0.05) for the comparison purposes. Networks were constructed for direct interactions between selected objects.

Expression quantitative loci (eQTL) identification was performed using data from the online GTEx portal (<https://www.gtexportal.org/home/>) to determine whether variants at 10^-7^ threshold for phenotype were eQTL in male and female-specific broad MDD GWAS, focusing on the central nervous system datasets (amygdala, anterior cingulate cortex, caudate, cerebellar hemisphere, cerebellum, cortex, frontal cortex, hippocampus, hypothalamus, nucleus accumbens, putamen, spinal cord, substantia nigra). Transcription factor analysis of the genes mapped from SNPs from male- and female-specific broad MDD GWAS at a p-threshold 10^-5^ were performed using MetaCore™.

We utilized a drug-target network building tool called Drug Targetor (drugtargetor.com) (5) to establish the potential mechanisms by which antidepressants act in male vs. female specific MDD. This resource uses Summary-PrediXcan (a statistical tool that assesses the mediating effects of gene information from summary statistics of genetic association studies on phenotypes; S-PrediXcan) from GWAS databases and drug/target interactions to assess phenotype-informed drug-target networks. The GWAS used was DEPR01: Major depressive disorder (6). The analysis was set to nervous system, the drug class to antidepressants, and the connection type to bioactivity and gene expression. We selected the maximum number of drugs possible (1500) and 50 drug targets. The gene targets from this analysis were further assessed using the ‘compare gene list’ functions in MetaCore® to determine the relation of male- or female-specific MDD associated genes altered by antidepressant medications.

**Validation of the sex specific GWAS through polygenic risk scores**

We then aimed at comparing the predictive capacity of the sex-specific MDD polygenic risk scores for detecting broad depression. For the sake of this comparison, we aligned the sample size of the male, female and total GWASs to avoid a power bias. Therefore, we selected ten random subsamples of the UK Biobank participants: one with the sample size similar to the males (129k participants) and another with the sample size similar to the females (147k participants) and conducted GWAS analysis on each of them. In all subsamples we maintained the same proportion of cases/controls and males/females as in the full UK Biobank sample retained for the analysis (274,141 subjects). As in the main analysis, for every selected subsample we applied linear regression (BGENIE) to access effect of each SNP on the adjusted broad depression phenotype. Then, we used the GWAS results to calculate the polygenic risk scores at different p-value thresholds using PRSice software (7,8) for each subject of the test sample (N=65,285 UK Biobank subjects not originally included in the main GWAS). For the comparison of the predictive ability of the different PRSs, we pooled together the results of the logistic regressions (10 for the females and 10 for the male sample). We used Akaike information criterion (AIC) to compare different models. This method identifies the best-fit model as the one that explains the greatest amount of variation using the fewest possible independent variables. Lower AIC scores associate with better-fitting models.
