## Supplemental Tables for "A Sex-Specific Genome-Wide Association Study of Depression Phenotypes in UK Biobank"

**Supplementary Tables**

**Table S1 -** The study demographics for the case and control groups within the broad MDD UK Biobank phenotype.

**A**

| **Total** | **Total** | **Control** | **MDD broad** |
| --- | --- | --- | --- |
| Number of subjects | 274141 | 177152 | 96989 |
| Sex |  |  |  |
| Female | 53.40% | 47.50% | 64% |
| Male | 46.60% | 52.50% | 36% |
| Mean age at recruitment | 56.8 (7.98) | 57.1 (8.06) | 56.4 (7.80) |
| Townsend deprivation index | -1.61 (2.92) | -1.77 (2.82) | -1.31 (3.07) |
| MDD broad % | 35.40% |  |  |

**B**

| **Males** | **Total** | **Control** | **MDD broad** |
| --- | --- | --- | --- |
| Number of subjects | 127867 | 92944 | 34923 |
| Mean age at recruitment | 57.1 (8.08) | 57.2 (8.15) | 56.7 (7.9) |
| Townsend deprivation index | -1.57 (2.98) | -1.72 (2.88) | -1.17 ( 3.2) |
| MDD broad % | 27.30% |  |  |

**C**

| **Females** | **Total** | **Control** | **MDD broad** |
| --- | --- | --- | --- |
| Number of subjects | 146274 | 84208 | 62066 |
| Mean age at recruitment | 56.6 (7.88) | 56.9 (7.96) | 56.2 (7.74) |
| Townsend deprivation index | -1.64(2.87) | -1.82 (2.76) | -1.39 (2.99) |
| MDD broad % | 42.40% |  |  |

**Table S2 -** Estimates of the intercept, standard error and genomic inflation factor obtained from linkage disequilibrium score regression

| Broad MDD GWAS | Intercept | Standard Error | Genomic inflation |
| --- | --- | --- | --- |
| **Total** | 1.0085 | 0.0076 | 1.2932 |
| **Females** | 1.0104 | 0.0066 | 1.1811 |
| **Males** | 1.0025 | 0.0066 | 1.127 |

**Table S3 -** Baseline characteristics for test sample

|  | **Total** | **Control** | **MDD broad** |
| --- | --- | --- | --- |
| Number of subjects | 65285 | 42038 | 23247 |
| Sex |  |  |  |
| Female | 55.40% | 49.70% | 65.80% |
| Male | 44.60% | 50.30% | 34.20% |
| Mean age at recruitment | 57.1 (7.97) | 57.4 (8.03) | 56.6 (7.82) |
| Townsend deprivation index | -1.48 (2.95) | -1.65 (2.86) | -1.16 (3.09) |
| MDD broad % | 35.60% |  |  |
