## Supplemental figures for "A Sex-Specific Genome-Wide Association Study of Depression Phenotypes in UK Biobank"

### Slide 1
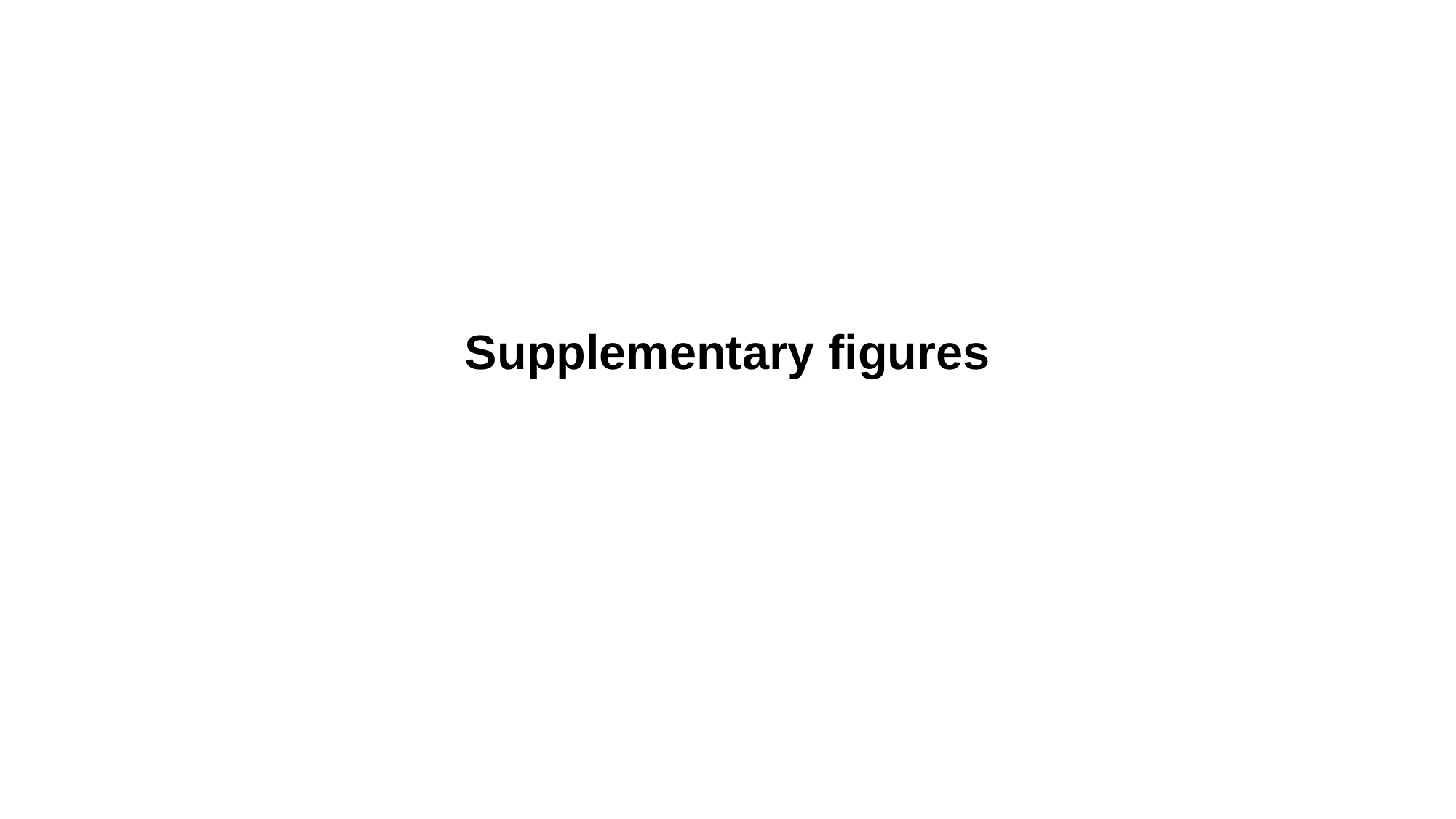

Supplementary figures

### Slide 2
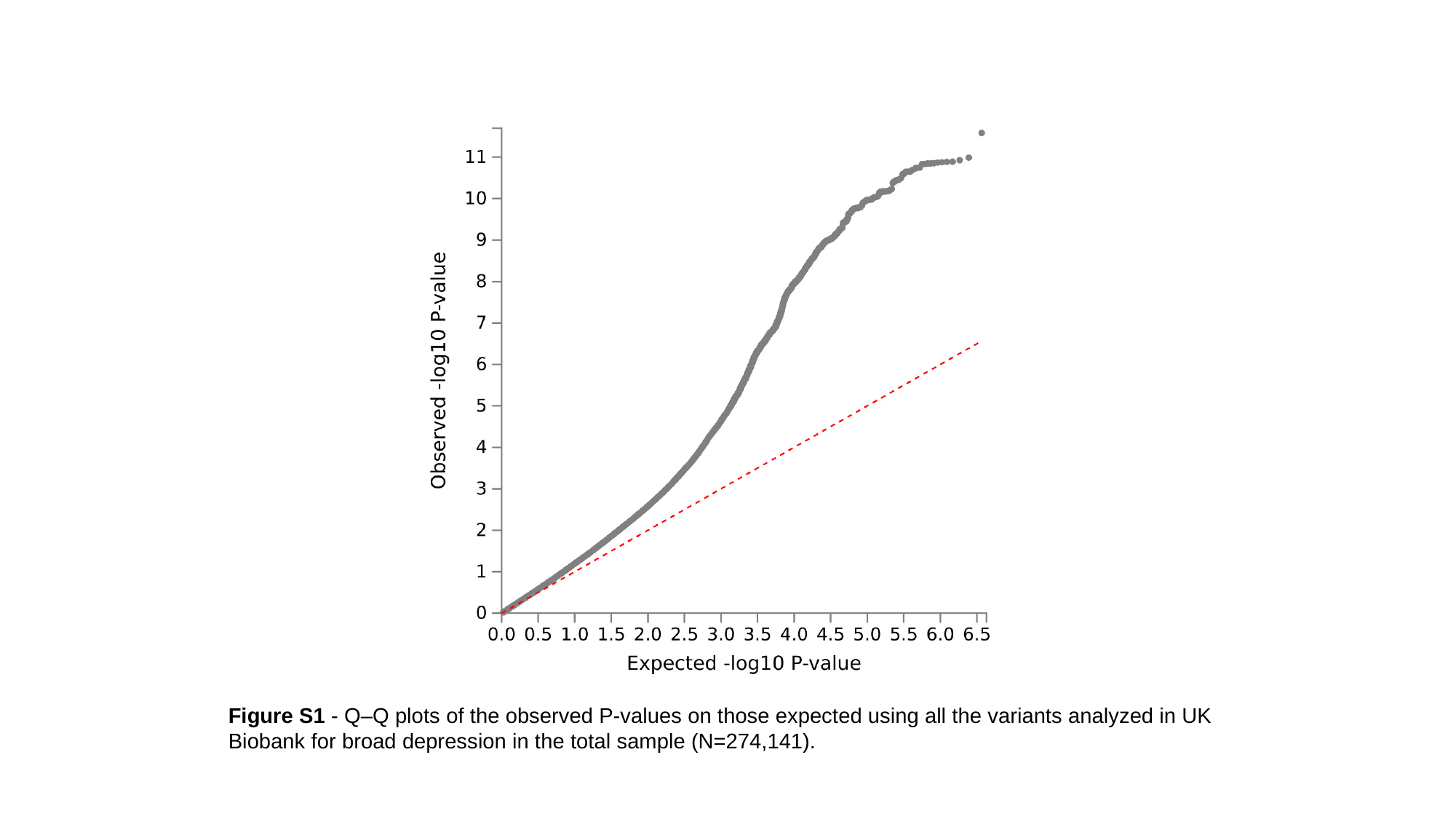

Figure S1 - Q–Q plots of the observed P-values on those expected using all the variants analyzed in UK Biobank for broad depression in the total sample (N=274,141).

### Slide 3
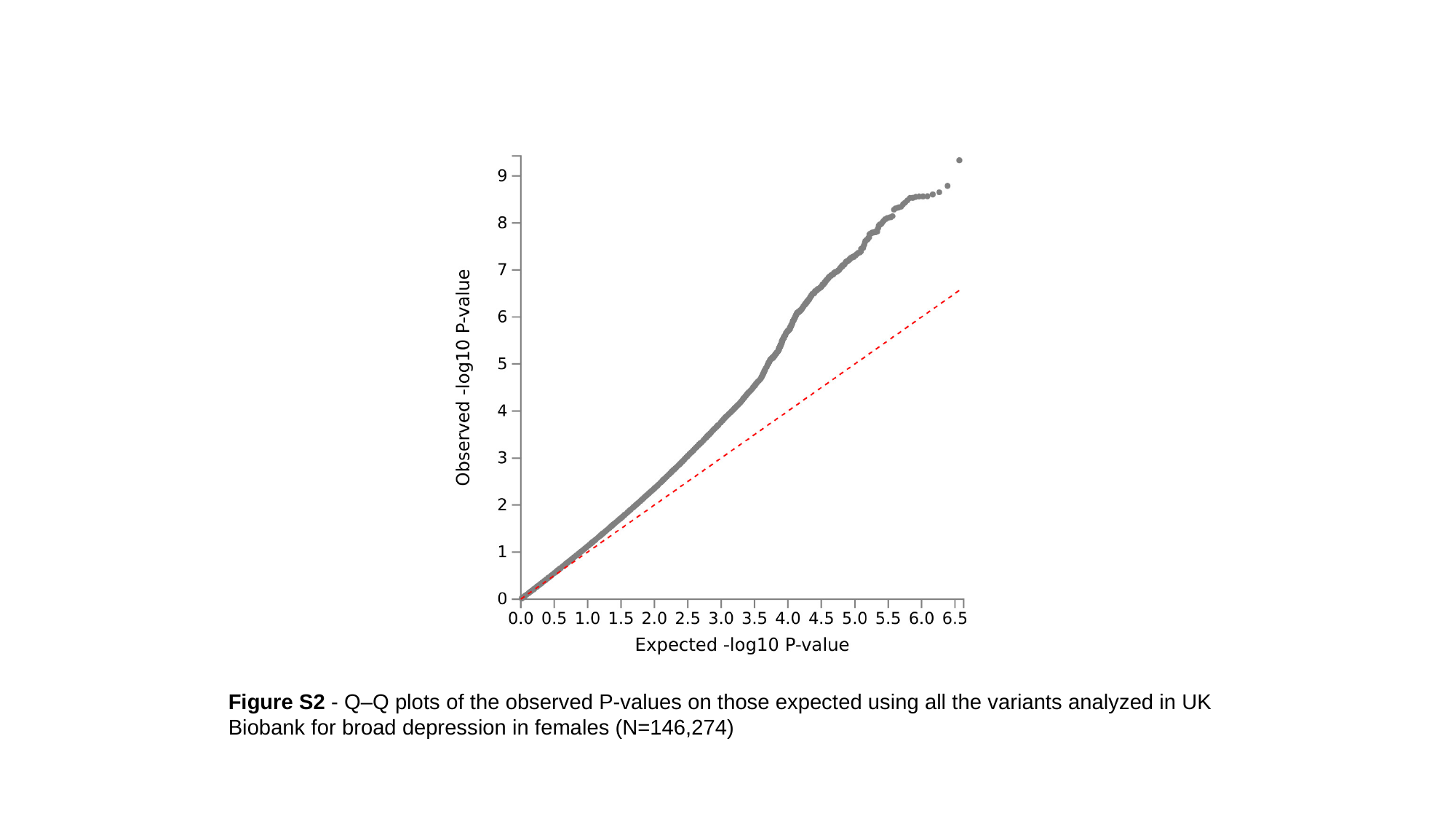

Figure S2 - Q–Q plots of the observed P-values on those expected using all the variants analyzed in UK Biobank for broad depression in females (N=146,274)

### Slide 4
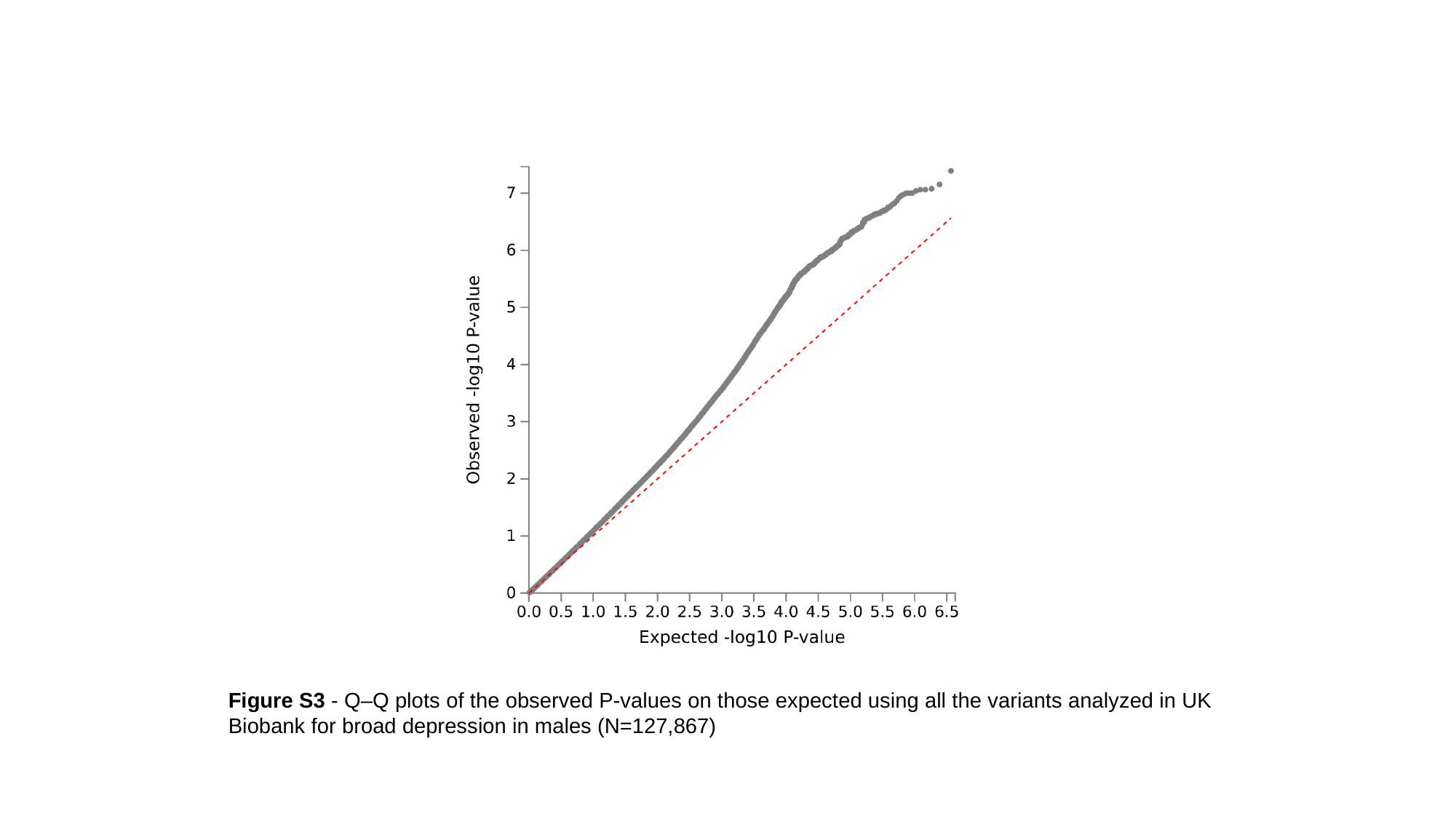

Figure S3 - Q–Q plots of the observed P-values on those expected using all the variants analyzed in UK Biobank for broad depression in males (N=127,867)

### Slide 5
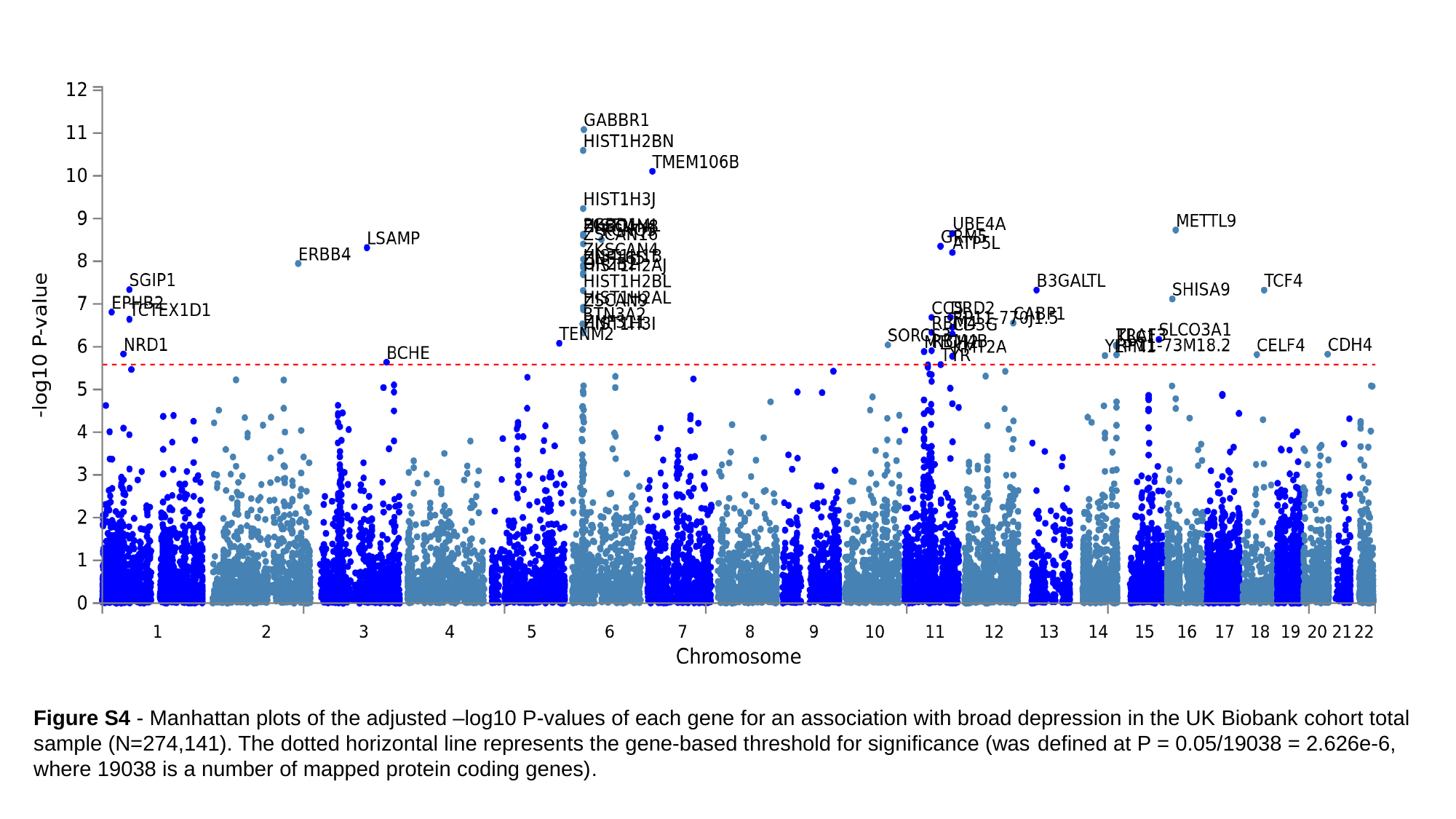

Figure S4 - Manhattan plots of the adjusted –log10 P-values of each gene for an association with broad depression in the UK Biobank cohort total sample (N=274,141). The dotted horizontal line represents the gene-based threshold for significance (was defined at P = 0.05/19038 = 2.626e-6, where 19038 is a number of mapped protein coding genes).

### Slide 6
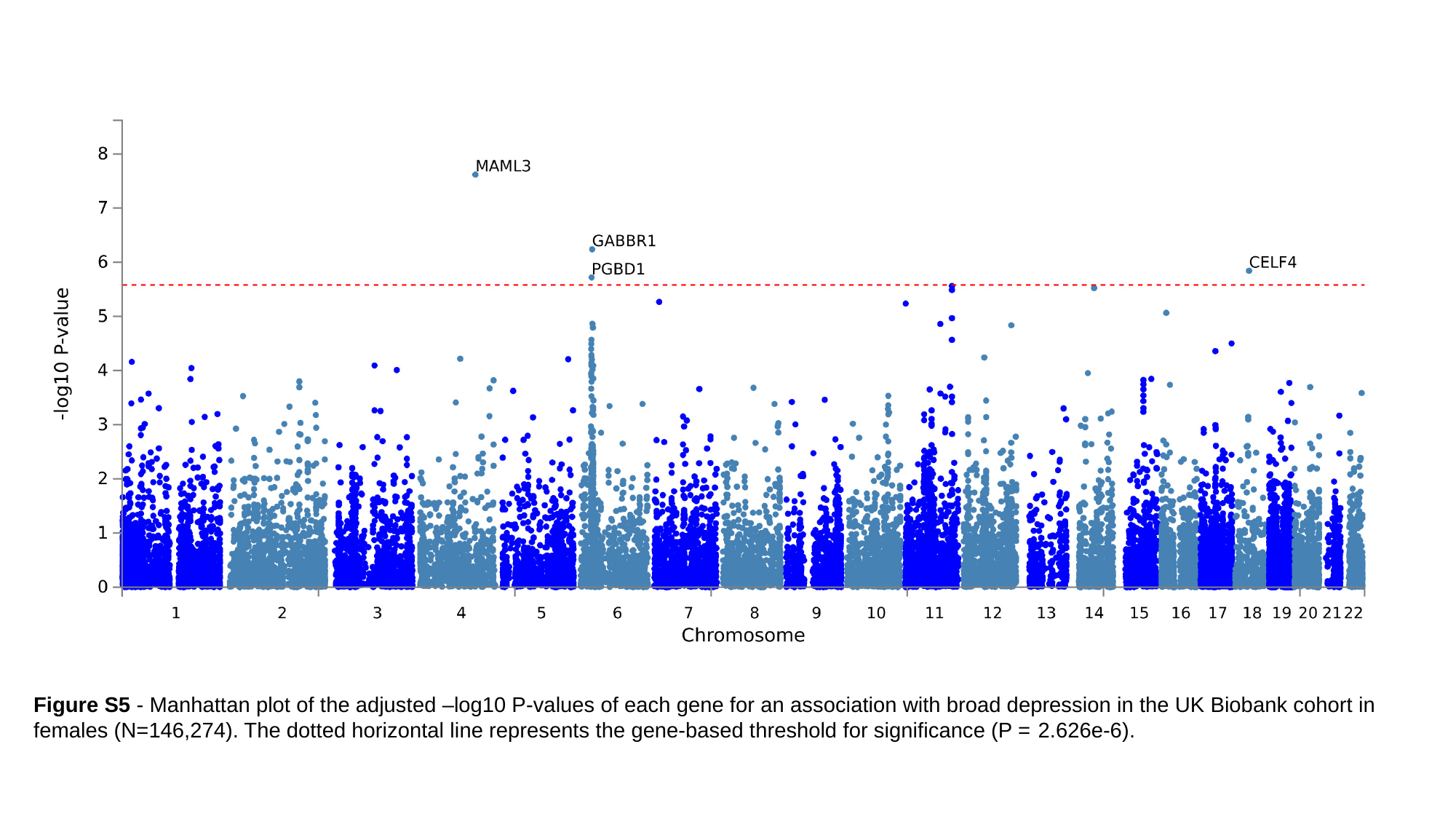

Figure S5 - Manhattan plot of the adjusted –log10 P-values of each gene for an association with broad depression in the UK Biobank cohort in females (N=146,274). The dotted horizontal line represents the gene-based threshold for significance (P = 2.626e-6).

### Slide 7
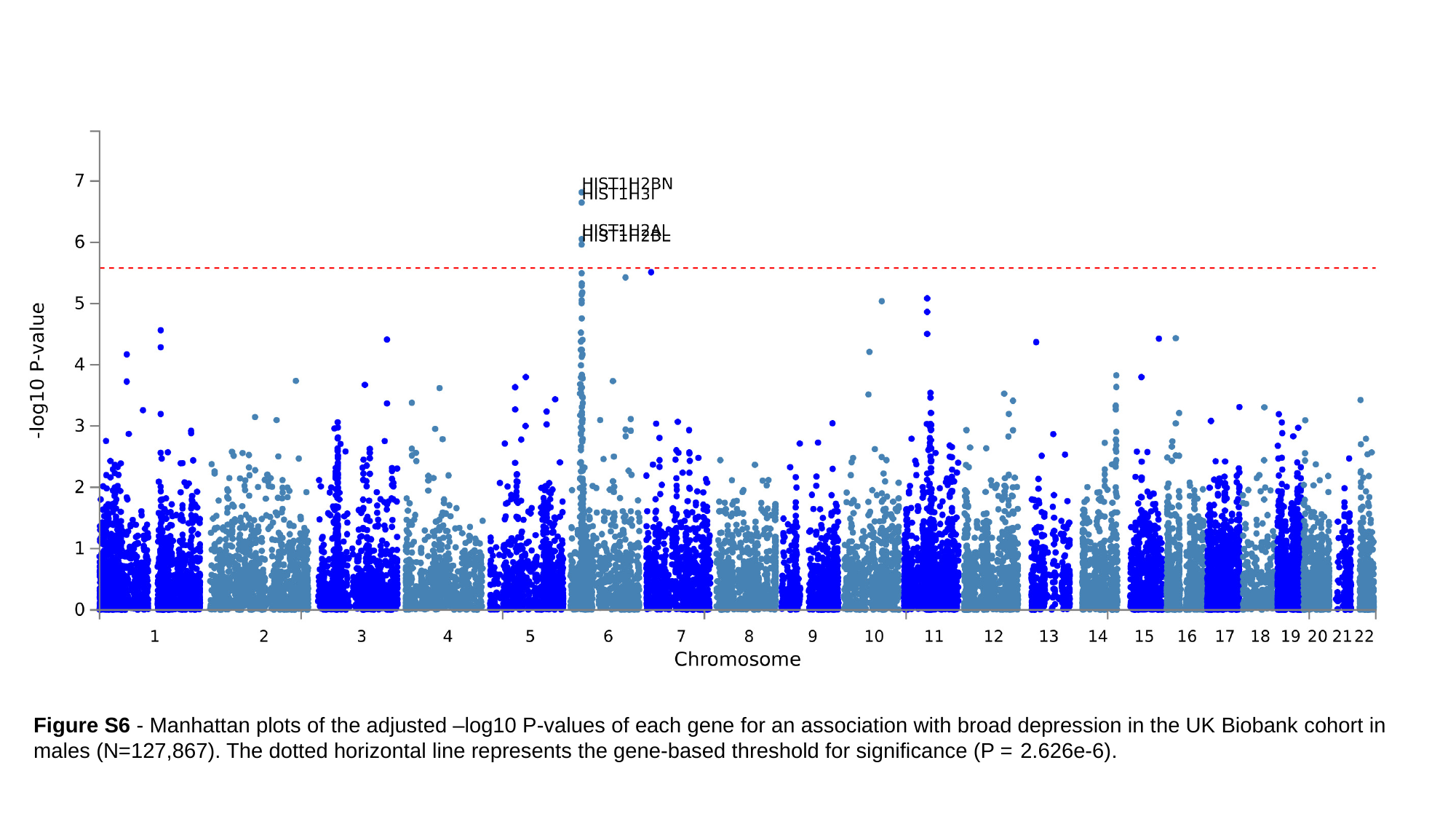

Figure S6 - Manhattan plots of the adjusted –log10 P-values of each gene for an association with broad depression in the UK Biobank cohort in males (N=127,867). The dotted horizontal line represents the gene-based threshold for significance (P = 2.626e-6).

### Slide 8
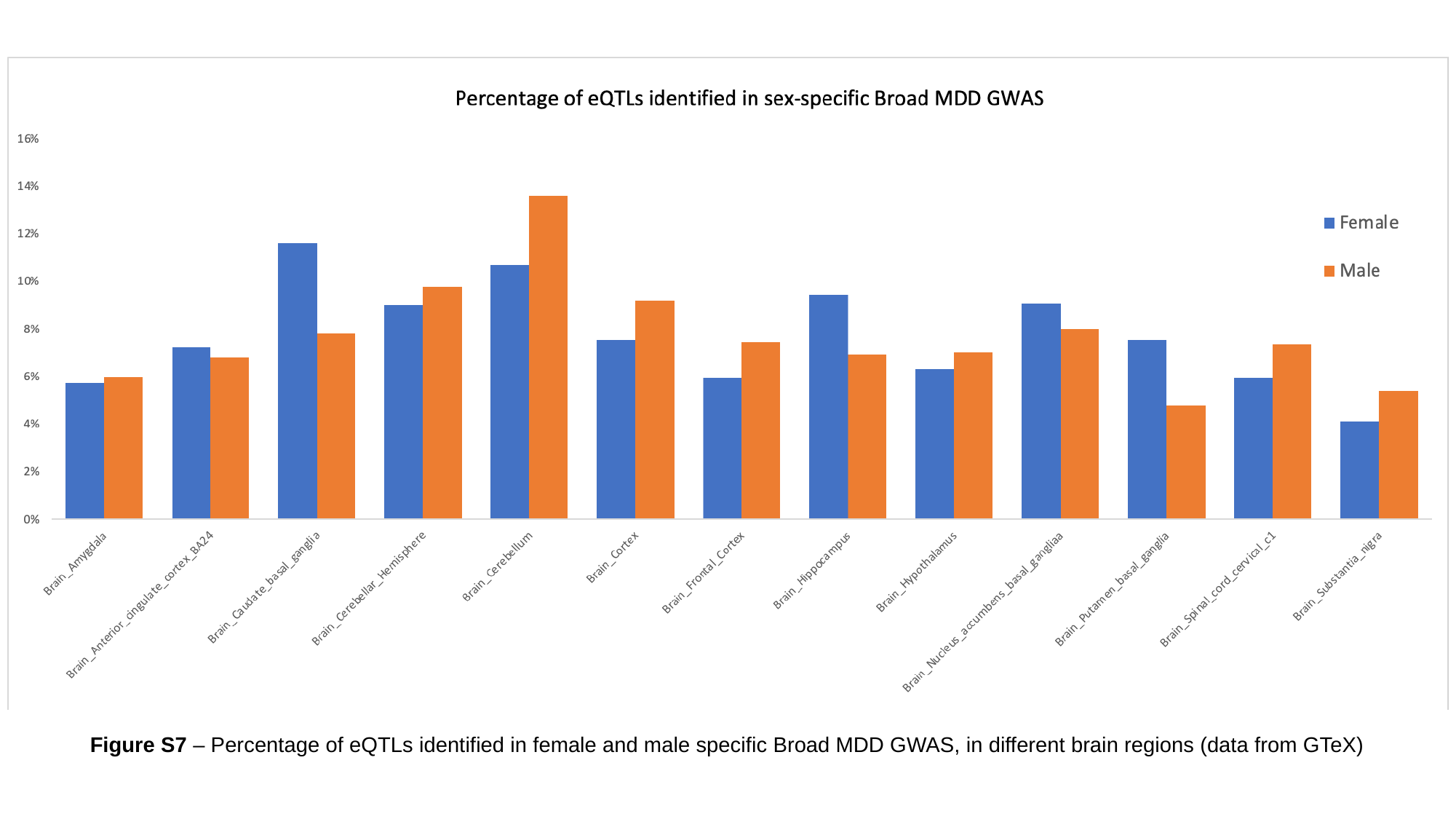

Figure S7 – Percentage of eQTLs identified in female and male specific Broad MDD GWAS, in different brain regions (data from GTeX)
